# Adaptive evolution and global spread of macrolide-resistant *Bordetella pertussis* during the post-pandemic pertussis resurgence

**DOI:** 10.1101/2025.07.15.25331551

**Authors:** Haodong Zhang, Zhihua Kang, Yingying Zhang, Yuanbin Yang, Huiwen Li, Naike Wang, Jiancheng Wang, Fuxing Jiao, Shanwei Tong, Bingsong Wang, Song Gao, Chengpei Ni, Xiaolu Shi, Shuang Wu, Simo Huang, Danni Bao, Qiushui He, Chuanqing Wang, Zhi Ruan, Pan Fu, Chao Yang

**Affiliations:** Laboratory of Microbiology, Department of Clinical Laboratory, Children’s Hospital of Fudan University, National Children’s Medical Center, Shanghai, China; Nosocomial Infection Control Department, Children’s Hospital of Fudan University, National Children’s Medical Center, Shanghai, China; Department of Laboratory Medicine, Shanghai East Hospital, School of Medicine, Tongji University, Shanghai, China; Department of Clinical Laboratory, Sir Run Run Shaw Hospital, Zhejiang University School of Medicine, Hangzhou, China; Department of Microbiological Testing, Ningbo Municipal Center for Disease Control and Prevention, Ningbo, China; Shanghai Institute of Materia Medica, Chinese Academy of Sciences, Shanghai, China; Pediatric Respiratory Department, Wuhu No.1 People’s Hospital, Wuhu, Anhui, China; The Affiliated Wuxi Center for Disease Control and Prevention of Nanjing Medical University, Wuxi Center for Disease Control and Prevention, Wuxi, China; Shenzhen Center for Disease Control and Prevention, Shenzhen, China; Department of Clinical Laboratory, Seventh Medical Center, PLA General Hospital, Beijing, China; Department of Clinical Laboratory, Sanmen People’s Hospital, Taizhou, China; Institute of Biomedicine, University of Turku, Turku, Finland; InFLAMES Research Flagship Centre, University of Turku, Turku, Finland; European Union (EU) Reference Laboratory for Diphtheria & Pertussis (EURL-PH-DIPE), Turku, Finland; Key Laboratory of Precision Medicine in Diagnosis and Monitoring Research of Zhejiang Province, Hangzhou, China; Zhejiang Provincial Engineering Research Center of Innovative Instruments for Precise Pathogen Detection, Hangzhou, China

## Abstract

Pertussis resurgence following the COVID-19 pandemic remains poorly understood. Here, we integrate global surveillance data with 8,117 *Bordetella pertussis* genomes from 35 countries to investigate the role of pathogen evolution. We identify substantial shifts in *B. pertussis* populations in China and Australia, alongside marked changes in multiple European countries. In China, resurgence is driven by the rapid expansion of a single macrolide-resistant clone, MR-MT28. Elsewhere, resurgence involves diverse, pre-pandemic polyphyletic strains. Australia and Europe show convergent antigenic changes, including a decline in pertactin-deficient strains, a rise in *prn2* alleles, and rising macrolide resistance.

Notably, we detect post-pandemic international dissemination of MR-MT28 across four continents, with non-Chinese isolates belonging predominantly to a pertactin-deficient subclone. These findings reveal convergent and region-specific strain replacement, antigenic evolution and macrolide resistance, highlighting the key role of adaptive evolution in the global pertussis resurgence. The emergence and global spread of MR-MT28 underscore the urgent need for coordinated global surveillance.

## Introduction

Pertussis, a highly contagious respiratory disease primarily caused by *Bordetella pertussis*, declined globally following the introduction of whole-cell and acellular vaccines^1,2^. Despite sustained high vaccination coverage, many countries have observed a resurgence over the past two decades, with a marked increase post-COVID-19 pandemic^3–5^.

According to the World Health Organization (WHO), following a sharp decline during the COVID-19 pandemic (2020–2022), global pertussis cases rose substantially post-pandemic (2023–2024), exceeding 800,000 notifications worldwide in 2024 —over at least four times higher than the pre-pandemic average (**figure 1A**). In Asia, China and South Korea reported >10-fold and >20-fold increases, respectively^6–8^. Several European countries, including France, Czechia and Denmark, recorded historically high incidence^9–12^. Similar surges were observed in Australia^13^ and the United States^4^ (**figure 1B**).

**Figure 1.**
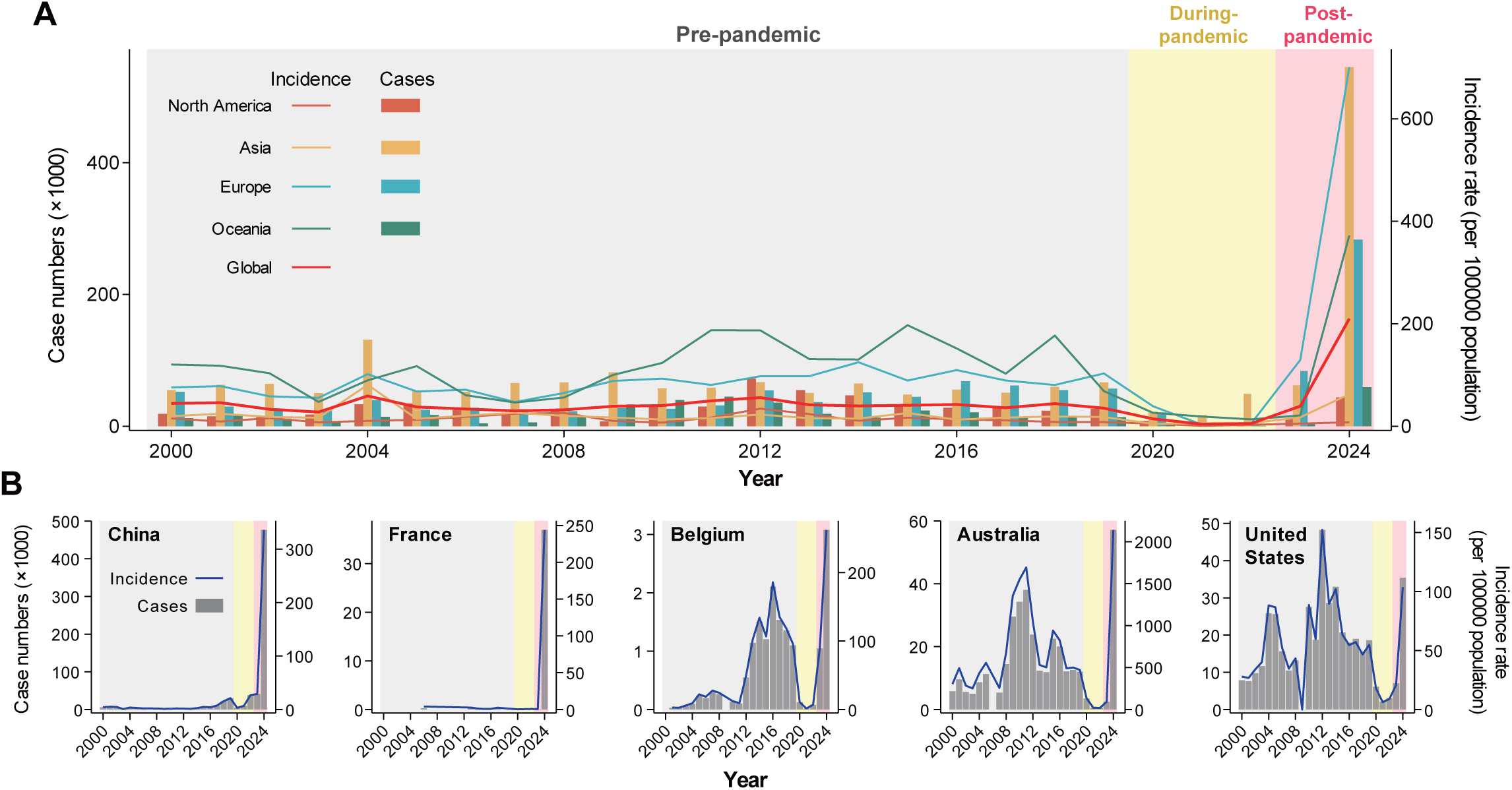
Global pertussis case numbers and incidence rates since 2000. (A) Trends in pertussis case numbers and incidence rates for the world and four continents. (B) Trends for four countries with more than ten *B. pertussis* genomes from pre-, during-, and post-pandemic periods. Incidence rates are shown per 100,000 population.

Multiple factors underlie this resurgence, including immunity debt from pandemic-related non-pharmaceutical interventions, waning vaccine-induced immunity, and adaptive evolution of *Bordetella pertussis*^2–7^. Over recent decades, *B. pertussis* has undergone notable genetic shifts, including the emergence and prevalence of strains with *ptxP3* alleles potentially associated with increased virulence^14^, and pertactin (PRN)-deficient variants, which may evade vaccine-induced immunity^15^. In parallel, the emergence of macrolide-resistant *B. pertussis* (MRBP), particularly its predominance in China, has raised additional concern regarding potential transmission and treatment failures^16–19^.

Of particular concern is the *ptxP3* MR-MT28 clone. It has been proposed as a major driver of China’s post-pandemic resurgence due to its combined features of vaccine escape and antimicrobial resistance^18,20^. Notably, this clone has been recently detected in France^11,12^ and Australia^13^, raising concerns of international dissemination. Although the post-pandemic resurgence of pertussis in China appears closely associated with the expansion of the MR- MT28 clone, integrated analyses of dynamics of circulating *B. pertussis* strains and rising incidence of pertussis before, during and after the pandemic at global level have been not performed.

In this study, we investigated the genomic evolution of *B. pertussis* in the context of the global post-pandemic resurgence. We found that lineage composition, vaccine antigen profiles, and MRBP prevalence underwent substantial shifts in several regions in parallel with the rising pertussis burden, providing evidence that adaptive evolution of *B. pertussis* is one key contributor to the recent global resurgence. Our analysis highlights that the MR-MT28 clone, which drove the post-pandemic resurgence in China, has now spread to other three continents, underscoring the urgent need for coordinated global surveillance.

## Results

### Global genomic diversity and population structure of *B. pertussis*

We analysed 8,117 high-quality (completeness >95%, contamination <5%) *B. pertussis* genomes, including 249 newly sequenced strains from six regions in China. These genomes spanned 35 countries across six continents. Four continents and five countries each had more than ten genomes from both pre-pandemic and during/post-pandemic periods, enabling temporal and geographical comparisons (**Fig. 2A**; Supplementary Fig. 1).

**Figure 2.**
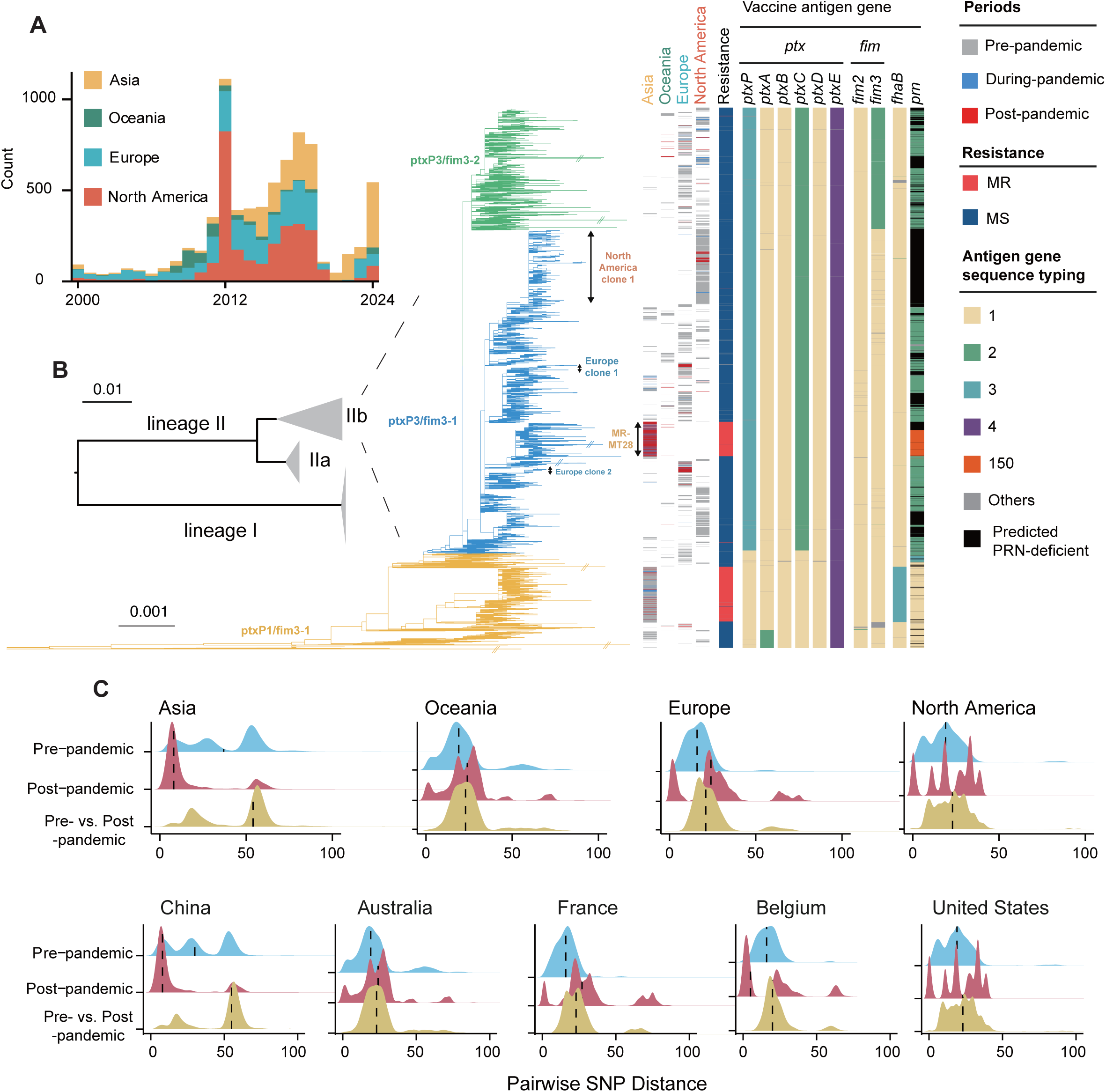
Global population structure and resistance and antigenic profiles of *B. pertussis*. (A) Annual number of high-quality genomes from four continents. (B) Maximum-likelihood phylogenetic tree of global isolates annotated with geographic location and macrolide resistance and vaccine antigen profiles. (C) Pairwise core-genome SNP distance distributions within and between time periods (pre-, during-, and post-pandemic) for each continent and representative countries.

A core-genome SNP-based phylogenetic tree was constructed to characterise the global population structure of *B. pertussis* (**Fig. 2B**). The phylogentic tree revealed that 99.6% (8087/8117) of isolates belonged to lineage IIb^30^, which could be further divided into three major sublineages based on *ptxP* and *fim3* genotypes: *ptxP1/fim3-1* (17%, n=1364), *ptxP3/fim3-1* (60%, n=4812), and *ptxP3/fim3-2* (22%, n=1816). The difference between *fim3- 1* and *fim3-2* alleles is a point mutation of C260A causing an amino acid change (Ala87Glu), and the difference between *ptxP1* and *ptxP3* is a point mutation of C to A in promoter of pertussis toxin gene.

The MRBP-associated A2047G mutation in 23S rRNA gene were detected in 59% (811/1364) of *ptxP1/fim3-1*, 11% (519/4812) of *ptxP3/fim3-1* and 0.3% (5/1817) of *ptxP3/fim3-2* strains. Nearly all (99%, 513/519) *ptxP3/fim3-1* MRBP strains belonged to the MR-MT28 clone (**Fig. 2B**), which was predominantly identified in China^20^.

### Lineage dynamics and variation in vaccine antigen and antibiotic resistance profiles

Post-pandemic increases in pertussis were accompanied by convergent and region-specific shifts in lineage composition, vaccine antigen profiles, and macrolide resistance (**Fig. 3**, Supplementary Fig. 2).

**Figure 3.**
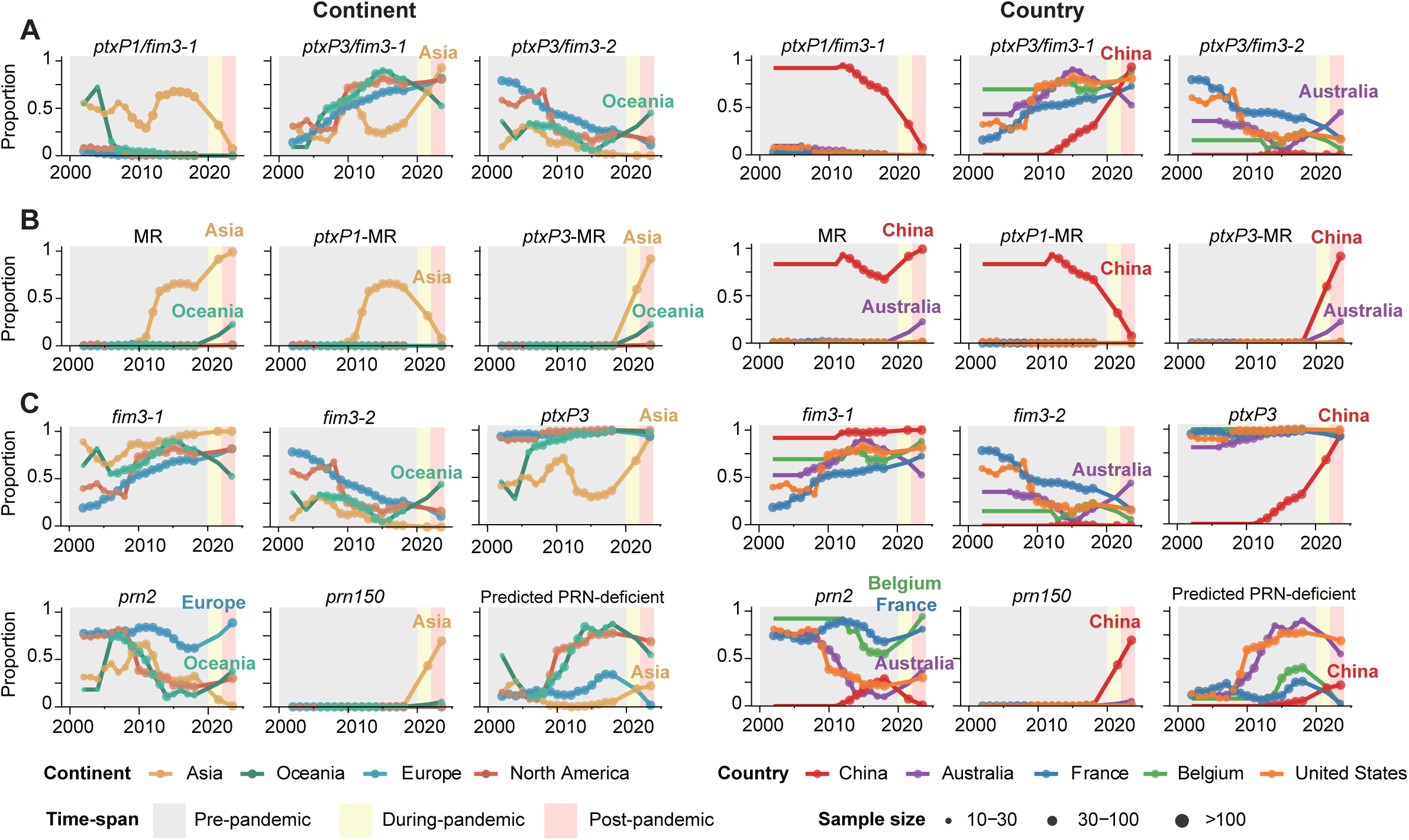
Temporal dynamics of *B. pertussis* lineages (A), macrolide resistance (B), and antigenic (C) profiles. Background colours indicate different time periods (pre-, during-, and post-pandemic). Lines show the proportion of strains. Point sizes indicate the number of genomes. Line and point colours represent each continent or country.

In Asia (n=2023), primarily represented by China (n=1645), the *ptxP3/fim3-1* sublineage expanded rapidly during and after the pandemic, alongside a marked rise in MRBP frequency and substantial shifts in antigen profiles (**Fig. 3**). This transition was driven by the emergence and rapid expansion of the MR-MT28 clone. MR-MT28 (n=513) carried either *ptxP3/fim3-1/prn150* (74%, 382/513) or *ptxP3/fim3-*1 with predicted PRN-deficiency (25%, 129/513) profiles, becoming the dominant clone (99%) post-pandemic in China (**Fig. 2B**, **Fig. 3**). This clonal replacement also shifted the MRBP composition, from a pre-pandemic predominance of *ptxP1/fim3-1* strains (87%, 774/891) to *ptxP3/fim3-1* (92%, 386/418) post-pandemic (**Fig. 3B**). In line with the dominance of a single clone, the genetic diversity of post-pandemic strains (median SNP distance: 8) was significantly decreased compared to that of pre-pandemic strains (median SNP distance: 31; **Fig. 2C**).

Similarly, in Oceania (n=489), mainly represented by Australia (n=445), substantial post-pandemic shifts were observed in sublineage distribution and antigen gene composition, accompanied by an increase in MRBP prevalence (**Fig. 3**). Based on this genomic analysis, MRBP prevalence notably reached 23% (9/40) post-pandemic—an unexpectedly high frequency given the previous rarity of MRBP outside China^19^, and this rise was also associated with the introduction of the MR-MT28 clone. Unlike Asia, changes in sublineage and antigen gene profiles were largely driven by the rise of *ptxP3/fim3-2* sublineage and *fim3- 2* and *prn2* alleles, along with a decreased frequency of predicted PRN-deficient strains (**Fig. 3**).

Post-pandemic strains in Oceania remained genetically diverse, with *ptxP3/fim3-2* and *ptxP3/fim3-1* sublineages present at 45% (18/40) and 53% (21/40), respectively. Accordingly, genetic diversity remained relatively stable post-pandemic. Phylogenetic analysis showed that close relatives of most post-pandemic strains were already circulating before the pandemic, with the exception of MR-MT28, suggesting that the resurgence in Oceania might be driven by the expansion of locally pre-existing lineages (**Fig. 2B**, **Fig. 3**).

In Europe (n=2595), mainly represented by France (n=1308) and Belgium (n=429), noticeable post-pandemic changes were observed in the vaccine antigen gene *prn*. As in Australia, the frequency of the *prn2* allele increased post-pandemic, while the frequency of predicted PRN-deficient strains decreased (**Fig. 3**). One MR-MT28 strain was detected in France in 2024.

Overall genetic diversity in Europe did not significantly decline post-pandemic, although two dominant clones (Europe clone 1 and 2, accounting for 65% of post-pandemic strains) were identified (**Fig. 2B**). Both clones were already present pre-pandemic, indicating continued transmission of locally endemic strains. However, this varied by country—for instance, post-pandemic strains in Belgium showed significantly reduced diversity, with 70% (64/91) belonging to Europe clone 2 (**Fig. 2B,C**).

In North America (n=2,832), primarily represented by the United States (n=2,798), strain dynamics remained relatively stable. The North America clone 1, prevalent pre-pandemic, remained dominant post-pandemic (67%, 69/103), continuing to drive endemic circulation (**Fig. 2B**). One MR-MT28 strain was identified in the United States in 2024.

### Evolutionary dynamics and global spread of the MR-MT28 clone

The MR-MT28 clone was detected in four countries (China, Australia, France, and the United States) across four continents post-pandemic (**Fig. 4A, B**), indicating its potential for global dissemination. We therefore investigated its evolutionary origin and patterns of international spread.

**Figure 4.**
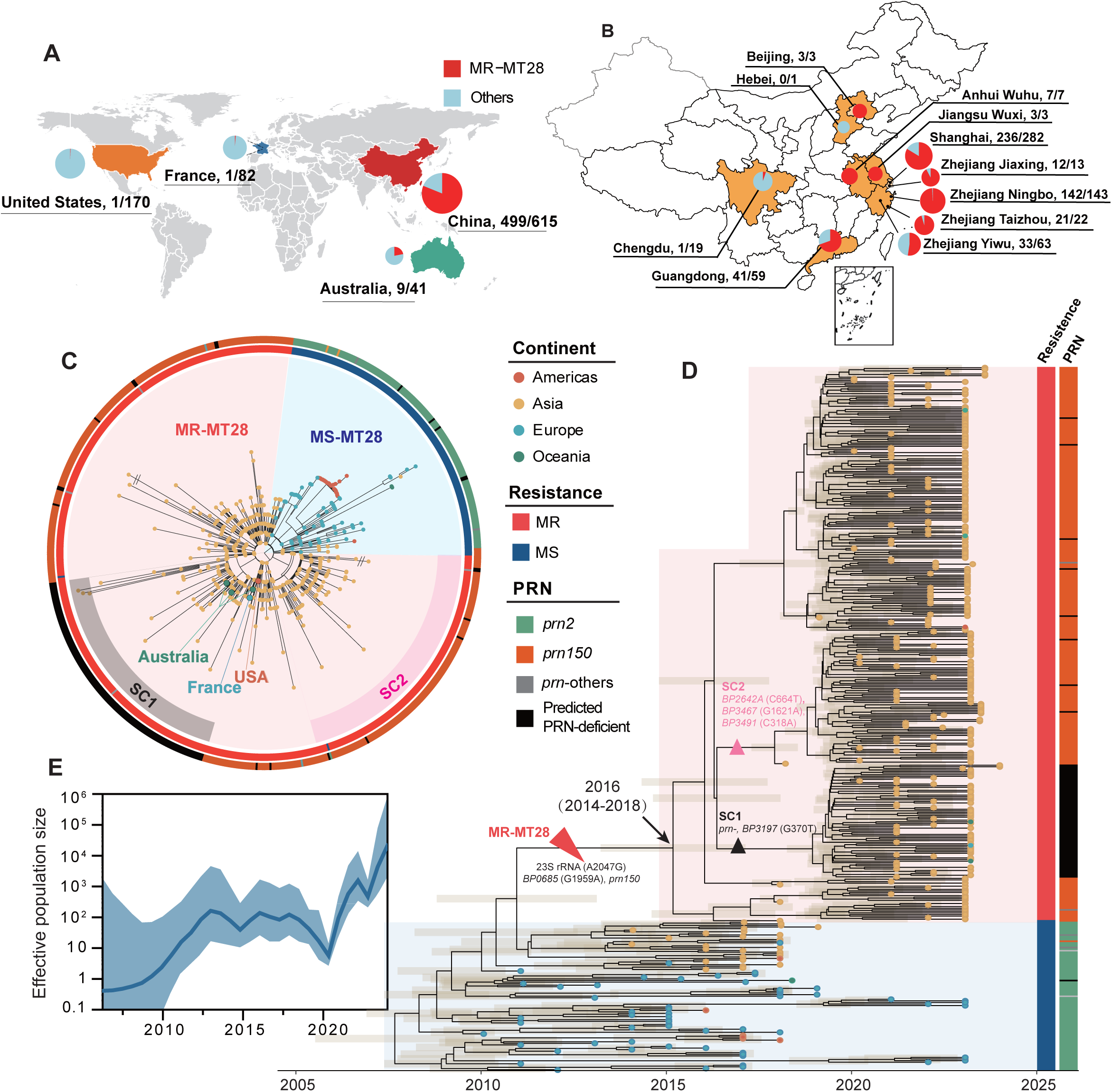
Evolutionary dynamics and global dissemination of the MR-MT28 clone. (A, B) Geographic distribution of MR-MT28 strains post-pandemic (2023–2024) globally and within China. Pie charts indicate the regional proportion of MR-MT28. (C) Maximum-likelihood phylogeny of the MT28 clone. Tip colours represent country of origin. Outer rings denote macrolide resistance (MR or MS) and *prn* allele profiles. (D) Time-calibrated phylogeny of the MT28 clone with resistance status and *prn* allele profiles. Key time points, genetic events, and 95% confidence intervals are indicated by arrows, triangles, and shaded orange rectangles, respectively. (E) Temporal changes in the effective population size of the MT28 clone inferred using Bayesian Skygrid analysis.

Phylogenetic analysis suggested that MR-MT28 evolved from the MS-MT28 lineage, which was primarily identified in Europe and North America pre-pandemic (**Fig. 4C, D**). Although MR-MT28 has become dominant in eastern China post-pandemic, it remains at relatively low frequency in western regions (**Fig. 4B**). Molecular clock analysis estimated the emergence of MR-MT28 around 2016 (95% CI: 2014–2018), most likely originating in China, where it now predominates. Consistent with the observed rise in MR-MT28 cases and its geographic expansion, its effective population size increased sharply post-pandemic—more than 1,000- fold in 2024 compared to 2021 (**Fig. 4E**).

Compared to its ancestor MS-MT28, the MR-MT28 clone harbours two defining mutations: the macrolide resistance-associated A2047G mutation in the 23S rRNA gene, and a synonymous G1959A mutation in *BP0685* (dehydrogenase/oxidase). The novel antigen allele *prn150*, found predominantly in MR-MT28, was also present in one MS-MT28 strain from Shanghai, China.

Two emerging subclones were identified within MR-MT28 (**Fig. 4C, D**). Subclone 1 (SC1), estimated to have emerged around 2019, is characterised by predicted PRN deficiency, a nonsynonymous mutation in *BP3197* (G287T; Arg96Leu; NAD(P)-dependent oxidoreductase), and, in a subset of strains, a mutation in *BP0026* (G370T; Ala124Ser; thiolase family protein). Notably, most (7/9) MR-MT28 strains detected in Australia and the single French isolate belonged to SC1, suggesting enhanced transmission potential, possibly associated with immune evasion.

Subclone 2 (SC2) harboured three distinct mutations: *BP2642A* (C664T; Pro222Ser; ABC transporter ATP-binding protein), *BP3467* (G1621A; Asp541Asn; AsmA family protein), and a synonymous mutation in *BP3491* (C318A; NAD(P)/FAD-dependent oxidoreductase).

Intriguingly, both subclones independently accumulated mutations in NAD(P)-dependent oxidoreductase genes (*BP3197* in SC1 and *BP3491* in SC2), suggesting convergent adaptation.

## Discussion

The global resurgence of pertussis appears to be multifactorial, shaped by both host-related and pathogen-related drivers^2–7^. Although the impact of host-relalted factors, such as non-pharmaceutical interventions and waning immunity, are difficult to quantify, genomic surveillance enables comprehensive characterisation of pathogen evolution. In this study, we systemically analyzed all available sequences of *B. pertussis* strains in the world and compared the results with incidences of pertussis before, during and after the COVID-19 pandemic. Our findings reveal substantial or noticeable post-pandemic shifts in *B. pertussis* lineage composition, vaccine antigen profiles, and macrolide resistance across three continents, with the most pronounced changes observed in China and Oceania.

In particular, we observed the rise of genotypes carrying mutations associated with antibiotic resistance, vaccine escape (PRN-deficient strains), and potential increases in virulence (e.g., *ptxP3* strains). The clearest evidence of such adaptive evolution was observed in China, where the *ptxP3* MR-MT28 clone, encoding a novel *prn150* allele, emerged as the dominant post-pandemic clone. This clone, capable of causing infections among older children and vaccinated individuals, was most likely the primary driver of the post-pandemic pertussis upsurge in China^18,20^.

Patterns of strain replacement and antigenic variation exhibited both convergent and divergent trends across continents. First, increased MRBP prevalence and higher frequencies of the *ptxP3* and *prn150* alleles were observed in both China and Australia post-pandemic, largely driven by the expansion of the MR-MT28 clone. Second, PRN-deficient strains declined in prevalence in Europe, Australia, and the United States after the pandemic, accompanied by a concurrent rise in the *prn2* allele^31^. Third, the *fim3-1* allele, already predominant before the pandemic, further increased in frequency in China, Europe, and the United States post-pandemic.

Region-specific trends revealed distinct adaptive trajectories. In China, the post-pandemic resurgence was driven by the clonal expansion of MR-MT28, in contrast to the polyphyletic strain composition observed in other regions. Notably, the prevalence of PRN-deficient strains uniquely increased in China post-pandemic, despite being rarely reported pre-pandemic and declining in frequency in many other countries^11,32,33^. This suggests that PRN deficiency may confer context- and time-dependent advantages, potentially shaped by regional vaccine formulations, the duration of vaccine use, and extent of exposure to pertussis in the population. In Australia, the frequency of the *fim3-2* allele uniquely rose post-pandemic. This allele had shown a global increase beginning in 1996, coinciding with the transition from whole-cell to acellular vaccines^30^. However, its global frequency has gradually declined since 2020 for reasons that remain unclear. The *fim3-2* allele has also been reported to be associated with enhanced biofilm formation^34^. Together, these findings may reflect region-specific selective pressures and potential local adaptation in China and Australia.

Despite the long-standing predominance of *ptxP1* MRBP strains in China for over a decade, they remain rarely detected elsewhere^16–19^. In contrast, *ptxP3* MR-MT28 clone has been identified across four continents, suggesting that it may possess distinctive features facilitating international spread. Indeed, the *ptxP3* strains appeared in Australia, Europe and the United States in 1990s and became prevalent since then. Our genomic analysis revealed a notable post-pandemic rise in MRBP prevalence in Australia, reaching 23% post-pandemic. This was further supported by targeted culture-independent sequencing screening, which estimated MRBP prevalence of 4.4%^13^. In addition to the MR-MT28 strain identified in this study from France, a recent French study showed that overall MRBP accounted for 2.8% (17/593) of the tests (culture and qPCR) in 2024. Phylogenetic analysis revealed that the 14 MRBP belong, together with isolates from China, to a cluster of 349 MRBP isolates. All 14 MRBP carried *ptxP3*, and 13 of them were PRN-deficient^11,12^. Moreover, MRBP strains have recently been identified in other European countries such as Finland^33^. Thus, the geographic range of MR-MT28 described here only represents a minimum estimate.

Most MR-MT28 strains detected in Australia and France were PRN-deficient, suggesting that PRN deficiency may confer additional advantages, such as enhanced vaccine escape potential. Together, these findings indicate that MR-MT28 represents a significant risk for global dissemination and underscore the urgent need for strengthened international surveillance, particularly targeting PRN-deficient MR-MT28 strains.

This study has several limitations. First, the publicly available genomic data used for contextualisation are subject to inherent sampling biases. Since the number of genomes available across countries and continents varies substantially, the results should be interpreted with caution. Second, recent genomic data from several regions, particularly the United States and parts of Europe, are limited or unavailable for comparison. Third, the number of isolates during the pandemic is very limited, which may affect our understanding of circulating strains during that period. Finally, vaccination programmes, vaccine types used, and non-pharmaceutical interventions varied across countries, and the resulting selection pressures in different populations may differ accordingly.

In conclusion, our study provides genomic evidence that the adaptive evolution of *B. pertussis*, through lineage replacement, antigenic variation, and antimicrobial resistance, has played a significant role in the post-pandemic resurgence of pertussis. The emergence and international dissemination of the MR-MT28 clone represent a growing public health challenge, emphasising the need for coordinated global surveillance and updated vaccine strategies.

## Methods

### Pertussis case data

Pertussis case numbers were obtained from the WHO website (https://immunizationdata.who.int/global/wiise-detail-page/pertussis-reported-cases-and-incidence). For countries or regions with incomplete WHO data, supplementary case counts were searched and sourced from national public health agencies, including the US Centers for Disease Control and Prevention (CDC) and the European Centre for Disease Prevention and Control (ECDC).

### Genome collection and sequencing

We analysed a total of 8,368 non-redundant *B. pertussis* genomes with unique Biosample accession numbers, comprising 8112 publicly available genomes and 249 newly sequenced isolates. Public genomes were retrieved from the NCBI Sequence Read Archive (SRA) and GenBank, with isolation year and country extracted from BioSample metadata. Newly sequenced isolates were collected from six regions in China: Shanghai (n=42), Zhejiang (n=165), Anhui (n=7), Jiangsu (n=3), Sichuan (n=19), and Guangdong (n=13). Whole-genome sequencing for newly sequenced isolates was performed on the Illumina NovaSeq platform. Newly generated sequencing data are available in the NCBI SRA under accession number PRJNA1288341.

Raw sequencing reads were quality-trimmed using Trimmomatic v0.39^21^. Genome assembly was conducted using the shovill v1.1.0 pipeline (https://github.com/tseemann/shovill).

Assembly quality was assessed using CheckM v1.1.3^22^. Only high-quality genomes (completeness >95%, contamination <5%) were retained in following analysis. Putative mutator strains indicated by abnormal tree branches were also removed from analysis.

After genome quality control, a total of 8117 high-quality genomes (Supplementary Table 1) were used in the analysis, includes isolates collected between 1900 and 2024 from 35 countries across six continents: Europe (16 countries), Asia (6), North America (5), Africa (4), Oceania (2), and South America (2).

### Phylogenetic and temporal analysis

Core-genome single nucleotide polymorphisms (SNPs) were identified using Snippy v4.6.0 (https://github.com/tseemann/snippy), based on raw reads or assemblies where reads were unavailable, with *B. pertussis* Tohama I (NC_002929.2) as the reference. SNPs within repetitive regions were excluded as previously described^23^. Maximum-likelihood phylogenies were constructed using FastTree v2.1.10^24^ (all strains) or RAxML-NG v1.0.1^25^ with the GTR+Γ model (MR-MT28 clone).

Time-resolved phylogenies and effective population size dynamics were inferred using BEAST v1.10.4^26^. We applied GTR+Γ substitution model and used isolation date (years) for tip date calibration. Six combinations of molecular clocks (strict and uncorrelated relaxed clock) and coalescent tree models (constant size and Bayesian Skyline and Skygrid) were tested and Skygrid model were the best fit based on path sampling and steppingstone sampling analysis. We ran ten independent Markov chain Monte Carlo chains for 30 million steps, sampling every 3,000 steps. The results of different runs were combined to generate the maximum clade credibility trees using TreeAnnotator v1.10.4. Effective population size dynamics was inferred using Tracer v1.7.1^27^.

### Vaccine antigen and macrolide resistance profiles

Vaccine antigen alleles were assigned by comparison with the curated *Bordetella* allele database on BIGSdb-Pasteur^28^. So far the only mechanism causing macrolide resistance has been the point mutation of A2047G in the 23S rRNA gene of *B. pertussis*. Therefore, MRBP strains carrying the A2047G mutation were detected using BLASTn. Strains were classified as PRN-deficient if they carried incomplete or disrupted *prn* coding sequences or promoters (BLASTn or TBLASTn coverage <99%), based on criteria defined by Lefrancq et al^29^.

## Data availability

Accession numbers for the publicly available genome sequences and associated metadata can be found in appendix and Zenodo (DOI:10.5281/zenodo.15900428). Newly sequenced genomes in this study are available from the National Centre for Biotechnology Information database under BioProject PRJNA1288341.

## Data Availability

All data produced in the present study are available upon reasonable request to the authors
All data produced in the present work are contained in the manuscript
Accession numbers for the publicly available genome sequences and associated metadata can be found in appendix and Zenodo (DOI:10.5281/zenodo.15900428). Newly sequenced genomes in this study are available from the National Centre for Biotechnology Information database under BioProject PRJNA1288341.

## Acknowledgments

This work was supported by grants from National Key Research and Development Program of China (2022YFC2304700), National Natural Science Foundation of China (82202567, 32270003), Youth Innovation Promotion Association, Chinese Academy of Sciences (2022278), and Shanghai Rising-Star Program (23QA1410500), Zhejiang Provincial Science and Technology Program for Disease Prevention and Control (2025JK139), Sigrid Juselius Foundation (240045) and Tampere Tuberculosis Foundation (26006205).

## Author contributions

P.F., C.W. and Y.C. contributed to the conception of this project. Z.R., Y.Z., P.F., Z.K., Y.Y. B.W., D.B., S.G., C.N., X.S., S.W., S.H. were responsible for collection of strains and metadata. H.Z., C.Y., H.L., N.W., J.W., F.J., S.T. performed bioinformatics analysis of whole genome sequence data. Y.C., P.F. and Q.H. prepared the manuscript. All authors contributed to the interpretation of results and critical review of the manuscript.

## Competing interests

We declare no competing interests.

## Notes

### Competing Interest Statement

The authors have declared no competing interest.

### Author Declarations

The isolates used in this study had been fully de-identified prior to use and cannot be linked back to the patients.

### Summary of Updates

We have update the version

