## Supplementaries for "Adaptive evolution and global spread of macrolide-resistant *Bordetella pertussis* during the post-pandemic pertussis resurgence"

**Supplementary appendix**

Supplementary figure 1. Geographic distribution of *B. pertussis* genomes pre-, during and post-pandemic periods. ........ 2

Supplementary figure 2: Temporal dynamics of *B. pertussis* macrolide susceptible (MS) strains, antigenic profiles (*fim2*, *fhaB*, *ptxP*, *ptxA-E*) and MR-MT28 clone. ... 3

Supplementary Table 1. Publicly available and newly sequenced Bordetella pertussis genome sequences included in this study. ........ 4

**Supplementary figure 1. Geographic distribution of *B. pertussis* genomes pre-, during and post-pandemic periods.**

The horizontal bar plot displays the number of high-quality genomes from 35 countries included in the global dataset, stratified by periods: pre-pandemic (before 2020; grey) and during/post-pandemic (2020-2024; red). The x-axis, representing the number of genomes, is shown on a logarithmic scale. For subsequent in-depth temporal and phylogenetic analyses, we selected countries with 10 or more available genomes from the pre- and during/post-pandemic period. The five countries that met this criterion were the United States, China, France, Australia, and Belgium.

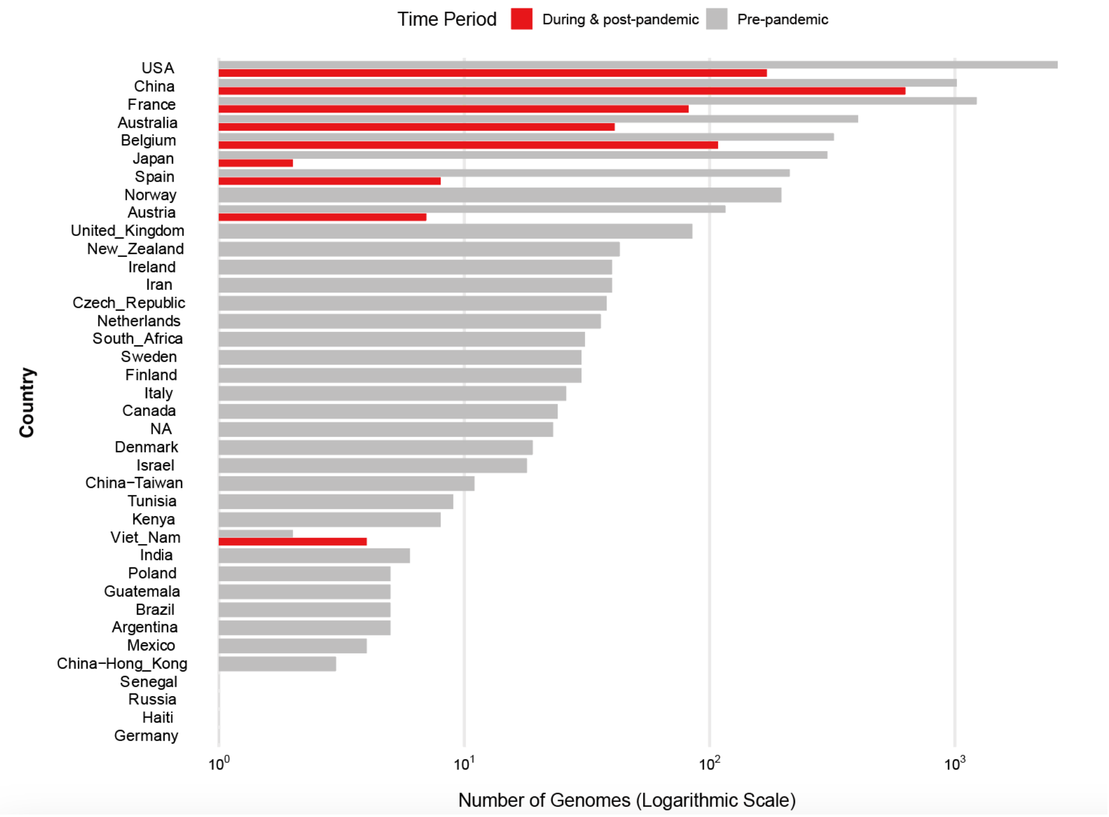

**Supplementary figure 2: Temporal dynamics of *B. pertussis* macrolide susceptible (MS) strains, antigenic profiles (*fim2*, *fhaB*, *ptxP*, *ptxA-E*) and MR-MT28 clone.**

Background colors indicate different time periods (pre-, during-, and post-pandemic). Lines show the proportion of strains; point sizes indicate the number of genomes; line and point colors represent each continent or country.

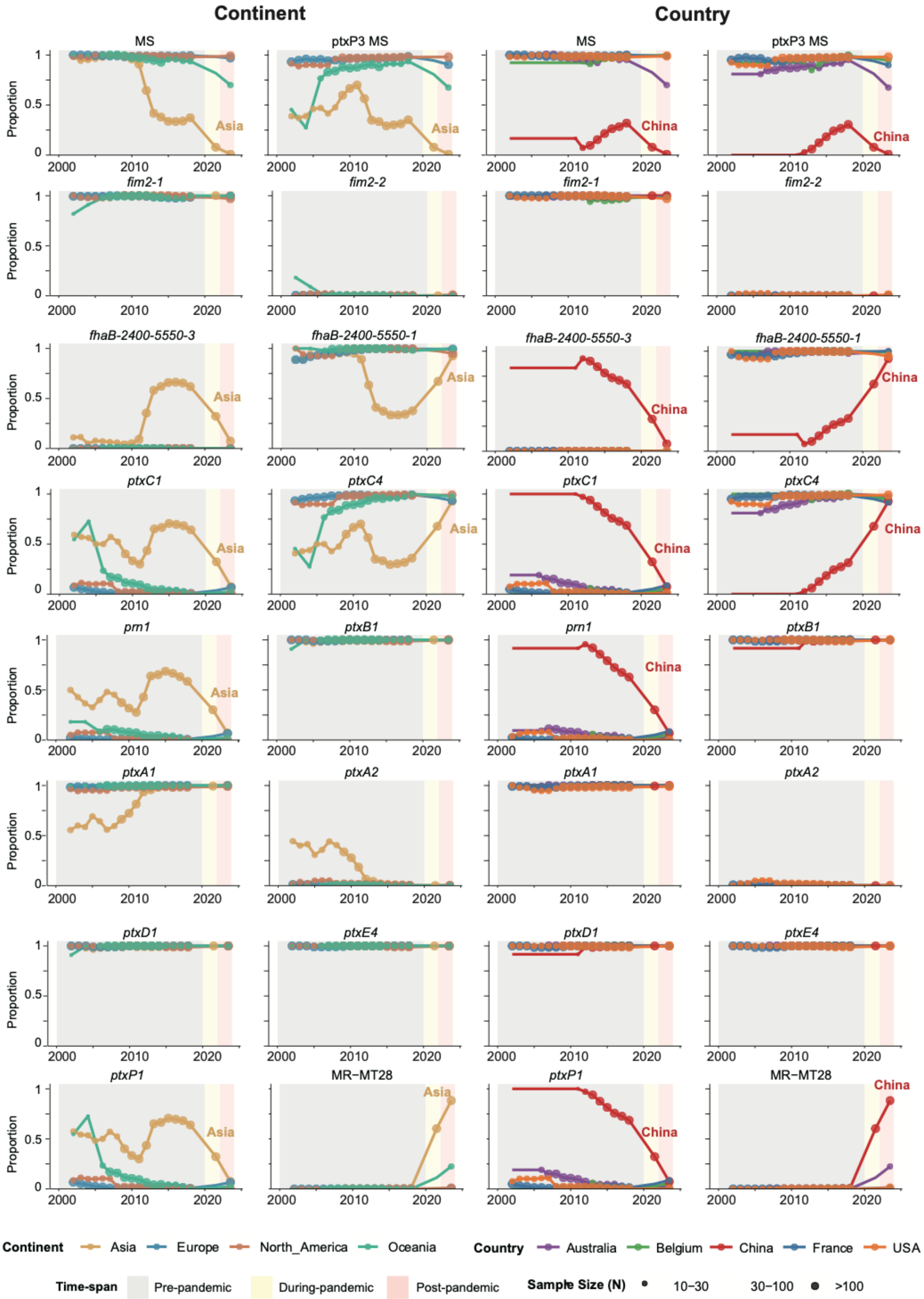

**Supplementary table 1. Publicly available and newly sequenced Bordetella pertussis genome sequences included in this study.**

The full dataset is available on Zenodo at [DOI:10.5281/zenodo.15900428].
